# Genomic Analysis of Human Brain Metastases Treated with Stereotactic Radiosurgery Under the Phase-II Clinical Trial (NCT03398694) Reveals DNA Damage Repair at the Peripheral Tumor Edge

**DOI:** 10.1101/2023.04.15.23288491

**Authors:** Jack M. Shireman, Quinn White, Namita Agrawal, Zijian Ni, Grace Chen, Lei Zhao, Nikita Gonugunta, Xiaohu Wang, Liam Mccarthy, Varshitha Kasulabada, Akshita Pattnaik, Atique U. Ahmed, James Miller, Charles Kulwin, Aaron Cohen-Gadol, Troy Payner, Chih-Ta Lin, Jesse J. Savage, Brandon Lane, Kevin Shiue, Aaron Kamer, Mitesh Shah, Gopal Iyer, Gordon Watson, Christina Kendziorski, Mahua Dey

## Abstract

Stereotactic Radiosurgery (SRS) is one of the leading treatment modalities for oligo brain metastasis (BM), however no comprehensive genomic data assessing the effect of radiation on BM in humans exist. Leveraging a unique opportunity, as part of the clinical trial (NCT03398694), we collected post-SRS, delivered via Gamma-knife or LINAC, tumor samples from core and peripheral-edges of the resected tumor to characterize the genomic effects of overall SRS as well as the SRS delivery modality. Using these rare patient samples, we show that SRS results in significant genomic changes at DNA and RNA levels throughout the tumor. Mutations and expression profiles of peripheral tumor samples indicated interaction with surrounding brain tissue as well as elevated DNA damage repair. Central samples show GSEA enrichment for cellular apoptosis while peripheral samples carried an increase in tumor suppressor mutations. There are significant differences in the transcriptomic profile at the periphery between Gamma-knife vs LINAC.

## Introduction

Paradoxically, the most commonly diagnosed brain tumor does not originate from the brain itself but is a consequence of systemic cancers^1, 2^. Clinically roughly 200,000 brain metastases (BM) are diagnosed annually, and they can be present in up to 30% of cases of primary malignancies such as lung, breast, melanoma. Currently, an effective treatment modality for local control of BM is radiotherapy, either targeted in the form of stereotactic radiosurgery (SRS), or untargeted in the form of whole brain radiation (WBRT)^2, 3^.

Given the significant amount of cognitive dysfunction associated with WBRT, SRS is emerging as the leading treatment modality for oligo-BMs (1 to 4 brain lesions), due to its ability to target radiation dose directly around the tumor, allowing for both the sparing of normal tissue as well as the delivery of a more concentrated radiation dose^2–5^. Two primary modalities of SRS include Gamma-Knife (GK) and linear accelerator (LINAC) based SRS and clinically they are considered comparable. However, a recent multi-institutional study comparing patients treated with GK vs LINAC for multiple brain metastasis showed higher incidence of radionecrosis in patients treated with GK compared to LINAC with similar overall survival^6^. These findings imply that the radiobiology of the two modalities of SRS delivery might be different and can be better understood at the genomic level. Currently SRS is being used as both a front-line therapy as well as in combination with surgical resection for symptomatic BM patients. Traditionally, SRS is delivered post-operatively to the resection cavity, however, several studies have hypothesized benefits that could be realized by pre-operative SRS^2–5, 7, 8^. Pre-operative SRS eliminates the need for surgical cavity margin expansion to be considered in the dosing scheme, while also ensuring that complications with surgical recovery do not impact the timing for delivery of SRS^9^. It has also been hypothesized that pre-operative SRS could prevent local leptomeningeal disease by dosing the tumors with focused radiotherapy, ideally limiting their proliferative capacity, prior to their disruption due to surgical resection^10^. Finally, data has also demonstrated that radiotherapy can be more effective on tumors which have intact vasculature as reactive oxygen species created by radiotherapy can be fully distributed within the tumor. Therefore, the act of surgical resection, and it’s inevitable disruption of tumor vasculature, may hinder the potential damage induced by SRS^11–13^. Although many of these radiobiological hypotheses have been tested in pre-clinical models of BMs, there are no studies analysis the radiobiological effect of SRS *in vivo* in human BMs to objectively guide clinical practice.

With the recent advancement in genomic analysis technology, a large body of research has accumulated attempting to uncover relevant driver mutations and genetic fingerprints of the BMs in general, with the goal of developing targeted therapies^14–20^. Notably lacking in the literature however is examination of the effects of clinical treatments, such as SRS and surgical resection, on both the BM itself as well its complex interaction with the surrounding microenvironment. Radiation therapy in general has been understudied in humans due to the limitations acquiring and analyzing clinically relevant radiated tissue samples^21–25^. Mouse studies have demonstrated effects such as an increase in corresponding immunotherapy responses to tumors (kindling effect) which depended only on a body part of the mouse being exposed to radiotherapy and was mediated by p53^26^. *In vitro* studies using both human and mouse cells have also implicated surrounding cells signaling, through cytokines and gap junctions, as critical players in the development and transfer of cytotoxic damage induced by radiation^24, 27, 28^. To address the current knowledge gaps in the literature regarding the optimal clinical sequence of SRS and surgery for the treatment of BMs and study the effect of SRS *in vivo* in human BMs at the genomic level we initiated clinical trial NCT03398694. The clinical goals of the trial are to evaluate the 6-month local control using pre-operative SRS for patients with 1-4 brain metastases and examine the rates of leptomeningeal disease (LMD) and radiation necrosis (RN). Scientifically, the trial leverages the unique opportunity of analyzing resected, radiated BMs allowing for direct examination of the genomic effects of SRS dosing on BMs in humans, producing an entirely novel dataset that can better inform clinical practice and basic science research.

In this study, we described the analysis of the first 34 patient’s SRS treated brain metastasis samples, obtained during surgical resection as part of the NCT03398694 trial. During surgical resection sections of the center of the tumor (relative to the SRS dosing field) as well as the peripheral edges were isolated and sent for genomic analysis using both whole exome sequencing (WES) and RNA sequencing (RNAseq) to comprehensively analyze the effect of SRS on tumor mutational landscape, local tumor and immune microenvironment, as well as in vivo genomic response to differing radiation dose and delivery. Analysis revealed that although similar radiation induced damage levels were seen between peripheral and central BM samples, damage locations varied randomly across the entire genetic landscape. Peripheral samples tended to enrich for DNA damage repair signatures while central samples showed enrichment for apoptotic damage signaling. Peripheral samples also demonstrated significant interaction with the surrounding brain microenvironment and propensity for mutations among tumor suppressor genes relative to central samples. Analysis of BM’s arising from different primary tumor locations revealed significant heterogeneity among transcriptional profiles indicating that site of origin of the BM has significant influence on its behavior in brain microenvironment. Finally, delivery of SRS with either gamma-knife (GK) or linear accelerator (LINAC) devices resulted in similar DNA damage locations but differential transcriptomic profiles across patients.

## Results

### Clinical trial details and patient characteristics

Phase-II clinical trial, NCT03398694, aims to enroll 50 patients with oligo-brain metastases. We performed genomic sequencing of the resected brain metastasis on the first 35 patients enrolled in the trial. Here we present the genomic analysis of 34 out of the 35 patients (Supplemental Table 1). One patient was dropped from the analysis because the final pathology was high-grade glioma. Out of the 34 patients, half of them were males and other half females. Age ranged from 36 to 85 years (median: 59) and except for one everyone had KPS of 70 or higher. Out of 34, 20 patients had previous diagnosis of cancer and 14 presented with precocious brain metastases. Primary cancer type included lung, breast, melanoma, renal, GI, and endometrial (Fig 1A). Patients who were enrolled in the trial received pre-operative SRS between 1-4 days before surgical resection of the BM. During surgical resection the center of the tumor along with the peripheral edges were sampled and stored for genomic analysis described in results and methods (Fig 1B). SRS dosing was based on RTOG 90-05^9^ and was delivered either by LINAC or Gamma Knife as single fraction. 4 out of 34 (11%) had treatment failure and local recurrence, 2 out of 34 (5.8%) had radionecrosis (RN) and 3 out of 34 (8.8%) developed leptomeningeal disease (LMD). Sufficient quality RNA and DNA was obtained from both peripheral and center matched tumors in 13 patients, with the other 21 trial patients lacking at least one component of RNA or DNA from a central or peripheral sample. In total all 34 patients had at least one component of usable sequencing data (Fig 1C), which was determined by extensive quality control metrics (Sup Fig 1A, B, C, & D). Overall, the analyzed dataset is balanced demographically to draw meaningful conclusions from the genomic studies.

**Figure 1:**
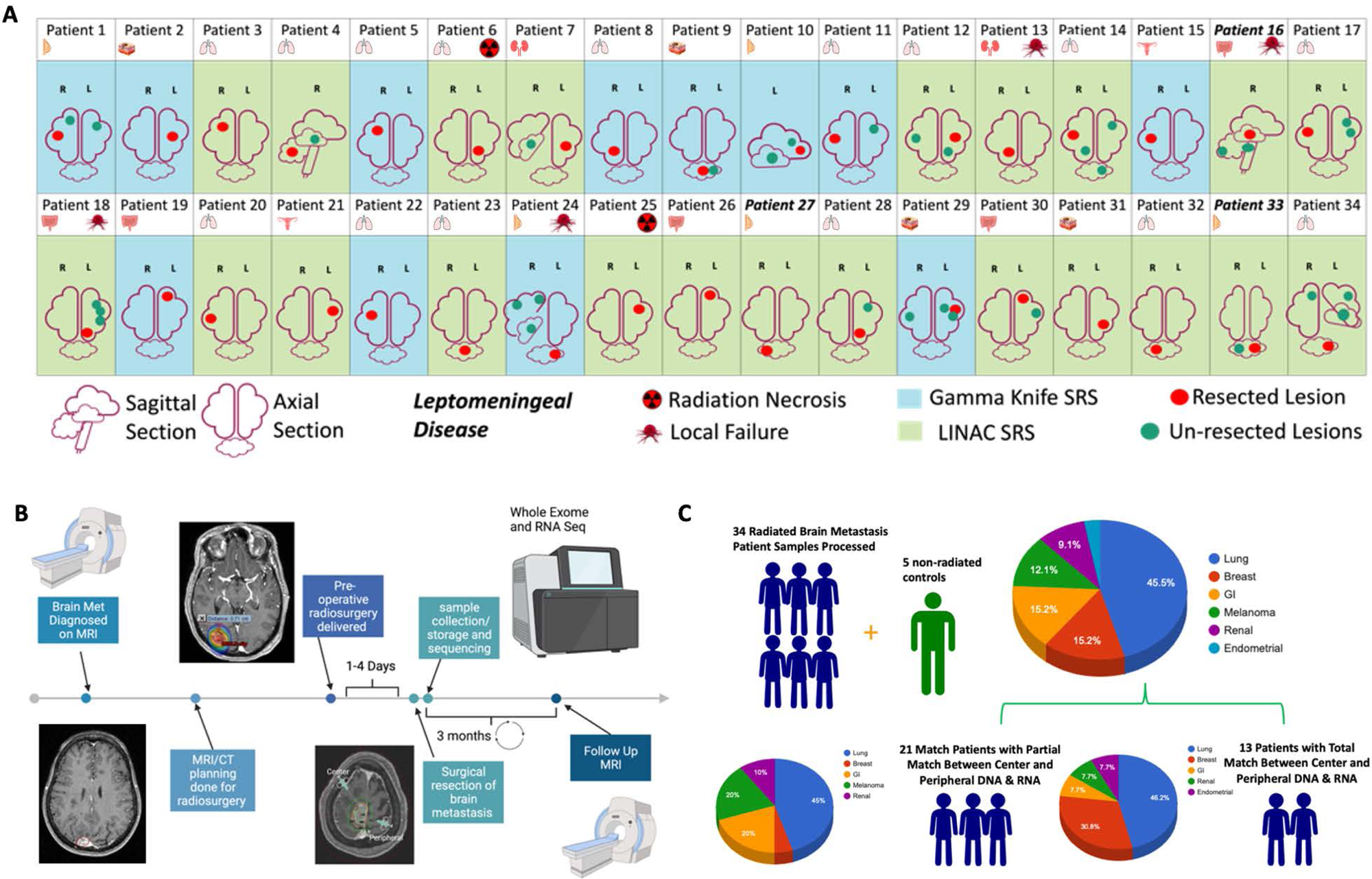
Clinical trial details and patient characteristics: **A)** Characteristics of the 34 patients enrolled in the clinical trial who contributed samples to the analyses contained within the manuscript. **B)** Schematic depicting diagnosis, radiosurgery timeline, and surgery and sample collection for clinical trial participants. **C)** Infographic depicting the primary tumor type of clinical trial participants and components of center or peripheral DNA and/or RNA contributed to the analyses.

### Pre-operative stereotactic radiosurgery incudes DNA damage leading to cellular changes in brain metastasis

The biological impact of radiation efficacy to halt spread, or in some case eliminate, brain metastases has been well documented clinically^2, 3, 29^, however there is very limited data available analyzing radiation induced damage at the genomic level within the human brain. Although the goal of radiation is to induce irreversible DNA damage in tumor cells resulting in tumor cell death, radiation can have other secondary effects on the tumor cells that can alter tumor cell biology *in vivo*. To accurately profile the DNA damage resulting from SRS to the BMs, as well as understand the possible functional outcomes of this cellular damage, we compared WES and RNA-seq done on irradiated BMs from the pre-op SRS clinical trial (Fig 2A). In these comparisons the central tumor samples were utilized to represent radiated tumors and control samples were separately obtained, type matched, lung brain metastases that received no radiation therapy. WES quantification of DNA damage (SNP, DNP, TNP, insertion, deletions) demonstrated a sharp increase in overall DNA damage in the tumors that received radiosurgery (Fig 2B, Sup Fig 2A) (non-radiated mean 841 marks vs Radiated mean 1849 marks, p<0.001), with the majority of damage coming from SNP’s within intronic regions (87.24%) (Sup Fig 2B). High impact variants annotated by SNPEFF^30^ demonstrated that between type matched non-small cell lung cancer (NSCLC) tumors, high impact deletions and insertions were more prevalent in radiated samples (Rad vs non-rad mean high impact variants detected: Deletion: 36 vs 18 p<0.01, Insertion: 17 vs 9 p<0.05) (Fig 2C). Total variant count per chromosome was visualized on a per sample basis indicating unique patterns of DNA damage was present among all samples (Fig 2D). Hierarchical clustering of high impact mutations on a per sample basis also demonstrated heterogeneity between patient samples (Sup Fig 2C). BCFtools^31^ vcfR^32^, and plotVCF were used to create gene summary plots for total radiated and non-radiated samples, revealing differential distribution of variants along chromosomes (Fig 2E). The C>T single base substitution (SBS), a hallmark of radiation damage^33, 34^, was the most prevalent in our sample (Fig 2F), while BM’s arising from melanoma primary tumors contained the highest fraction of SBS marks per radiated tumor type analyzed, likely due to the lesions propensity for prior UV induced radiation damage^22, 35–37^ (Fig 2G). Waterfall plots were created from gene ontology analysis of differentially expressed genes present in the radiosurgery cohort using ALLEZ^38^ (p <0.05 significance level with previous term exclusion). Results indicated DNA repair as the top enriched term along with other cellular growth processes such as ATP-dependent activity, centriole formation, and regulation of DNA replication (Fig 2H, Sup Fig 2D). DeSeq2^39^ was used for differential expression analysis between type matched tumors receiving radiosurgery or no radiosurgery 143 terms were enriched (α-threshold <0.01) (Fig 2I). Genes contributing to the global ALLEZ GSEA enrichment from terms double strand break and DNA repair genes were then isolated to visualize expression changes confirming upregulation among the radiated tumors across both LINAC and GK systems (Fig 2J & K). Finally, dimensionality reduction followed by UMAP based clustering was performed on samples revealing moderate clustering of samples from the same tumor (center vs periphery) but not across patients (Sup Fig 2E). These data illustrate that in humans SRS induces measurable and significant cellular damage and distress across a range of metastatic tumors from differing primary locations. The functional consequences of the induction of cellular damage seems to be enrichment for cellular repair which is not surprising given cancer cells have proven adept at proliferating despite higher than baseline levels of cellular damage or mutations^23, 40–44^.

**Figure 2:**
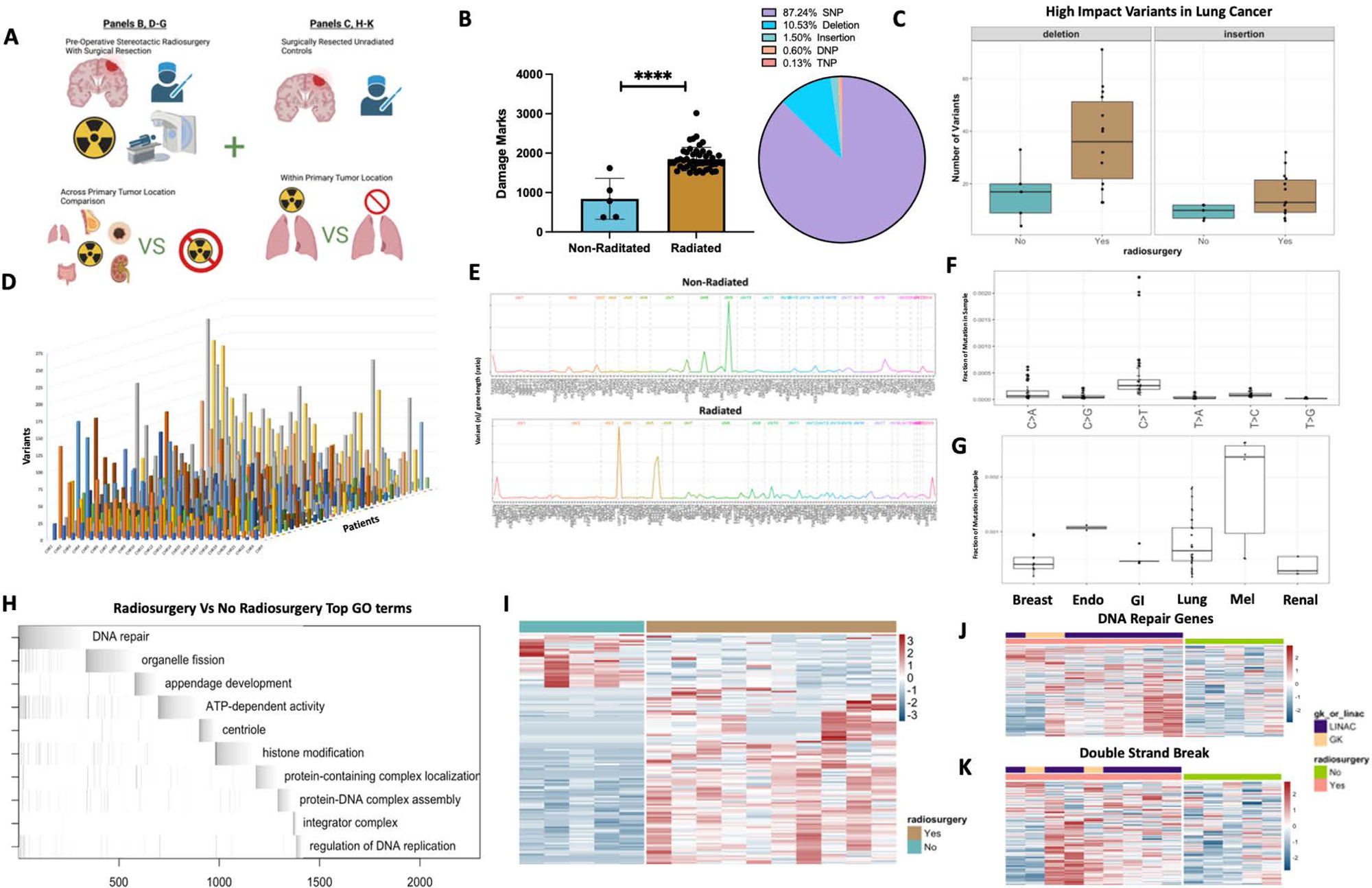
Pre-operative stereotactic radiosurgery induces DNA damage leading to cellular changes in brain metastasis: **A)** Schematic depicting analysis of samples with and without radiosurgery compared between common primary tumor locations (lung) or across primary tumor locations. **B)** Quantification of total damage marks and type of damage detected by variant calling between radiated and non-radiated samples. **C)** Comparison of high impact deletions or insertions among radiated and non-radiated lung cancer samples. **D)** 3D plot of all samples organized by chromosome (x), number of variants detected (y), and patient (z). **E)** Chromosome visualization plot of mutated genes across non-radiated and radiated samples. **F)** Single base substitutions present in radiated samples. **G)** Single base substitutions present grouped by primary tumor location across radiated samples. **H)** ALLEZ GSEA waterfall plot with previous term exclusion applied to genes differentially expressed among tumors metastasized from lung. **I)** Heatmap visualization of differentially expressed genes among non-radiated samples (left/blue) and radiated samples from tumors metastasized from lung (right/brown). **J)** Heatmap visualizing expression of genes associated with GO term “DNA Repair” among non-radiated and radiated tumors metastasized from lung. **K)** Heatmap visualizing expression of genes associated with GO term “Double Strand Break” among non-radiated and radiated tumors metastasized from lung. Comparison between samples done using students t-test (B), differential expression analysis was done within DESeq2 or ALLEZ using adjusted p-value < 0.05 (H, I, J, K). **** p< 0.0001

### Radiation dosing fall-off along peripheral edges of brain metastasis induces DNA damage repair and enrichment of cellular growth genes

The main hypothesized benefit of pre-operative SRS is to ensure that metastatic tumors receive the maximal conformed dose before surgery disrupts and disperses the tumor cells into the surrounding cortical tissue. Although SRS delivers a concentrated dose of radiation to the lesion, there is differential dosing between the center and periphery of the treated lesion^45–47^. The optimal dosing for treating a lesion strives to strike the balance between maximizing radiation induced cancer cell apoptosis (disease-control) and minimizing radiation induced brain necrosis (radionecrosis). With continued improvement in overall survival of BM patients with new targeted systemic therapies, optimal delivery of SRS to maximize disease-control and minimize radionecrosis remains a critical clinical consideration. Currently, there is no data in humans examining the comparative genomic effect of differential radiation dosing between center and periphery of a treated lesion. We hypothesized that the differential radiation dosing will have differential effect on the overall genomic signature of the center and periphery of a treated lesion which may explain clinical behavior such as treatment resistance or failure. To test this hypothesis, we examined surgical biopsies from the center, and peripheral edges, of BM’s using both WES and RNA sequencing (Fig 3A). Quantification of shared versus unique variants indicated most variants were common among center and peripheral samples, however, unique variants still comprised roughly 20% of the variants detected (Fig 3B & C). Classification of variant type with SNPEFF and COSMIC^48^ indicated similar numbers of low, modifier, moderate, and high impacts across center and peripheral samples while also demonstrating the majority of variants detected (∼2/3) were not present within the existing COSMIC database (Sup Fig 3A & B). Visualization of variants across chromosomal location between the center and peripheral samples shows globally very similar overlap patters with different variant location visible on an individual chromosome level (bottom vs top) (Fig 3D). Analysis of variants per gene across the samples demonstrated consistency within sample pairs but unique variation across primary tumor types (Fig 3D and Sup Fig 3C). DIFFUSE^49^ was utilized to perform isoform enrichment between center and peripheral samples allowing for a more in-depth transcriptional picture of the underlying biology. Isoform enrichment revealed significant transcriptional variation can exist within the same tumor sample (Fig 3F, Sup Fig 3D), something seen in high grade glioma biology^50–53^ but not yet described in the BM population. Gene ontology enrichment terms included leukocyte activation, extracellular matrix, and angiogenesis (Fig 3G). Furthermore, significant enrichment of DNA double strand break processing was enriched among peripheral samples while apoptotic signaling was enriched among center samples (Fig 3H), possibly resulting from SRS dose falloff due to the nature of radiation beam conforming. Peripheral samples also had higher numbers of mutations present with annotated tumor suppressor genes (Fig 3I), possibly increasing proliferative capacity of the cells within this region. To examine interactions with surrounding brain and immune cells FARDEEP^54^ devolution analysis was conducted which demonstrated peripheral sample enrichment for excitatory neuronal cells (Fig 3J) and immune invasion dominated by the macrophage and plasma cell lineage (Fig 3K). IHC of central and peripheral samples corroborated our in silico finding demonstrating robust CD45+ immune cell invasion (Fig 3L) (Lung: center vs peripheral CD45+ counts p< 0.05). These data demonstrate that the small difference in radiation dose received by peripheral edges of tumors compared to the center likely plays some role in allowing for more functional interaction with the surrounding microenvironment, upregulated DNA damage repair, as well as increased proliferation capacity.

**Figure 3:**
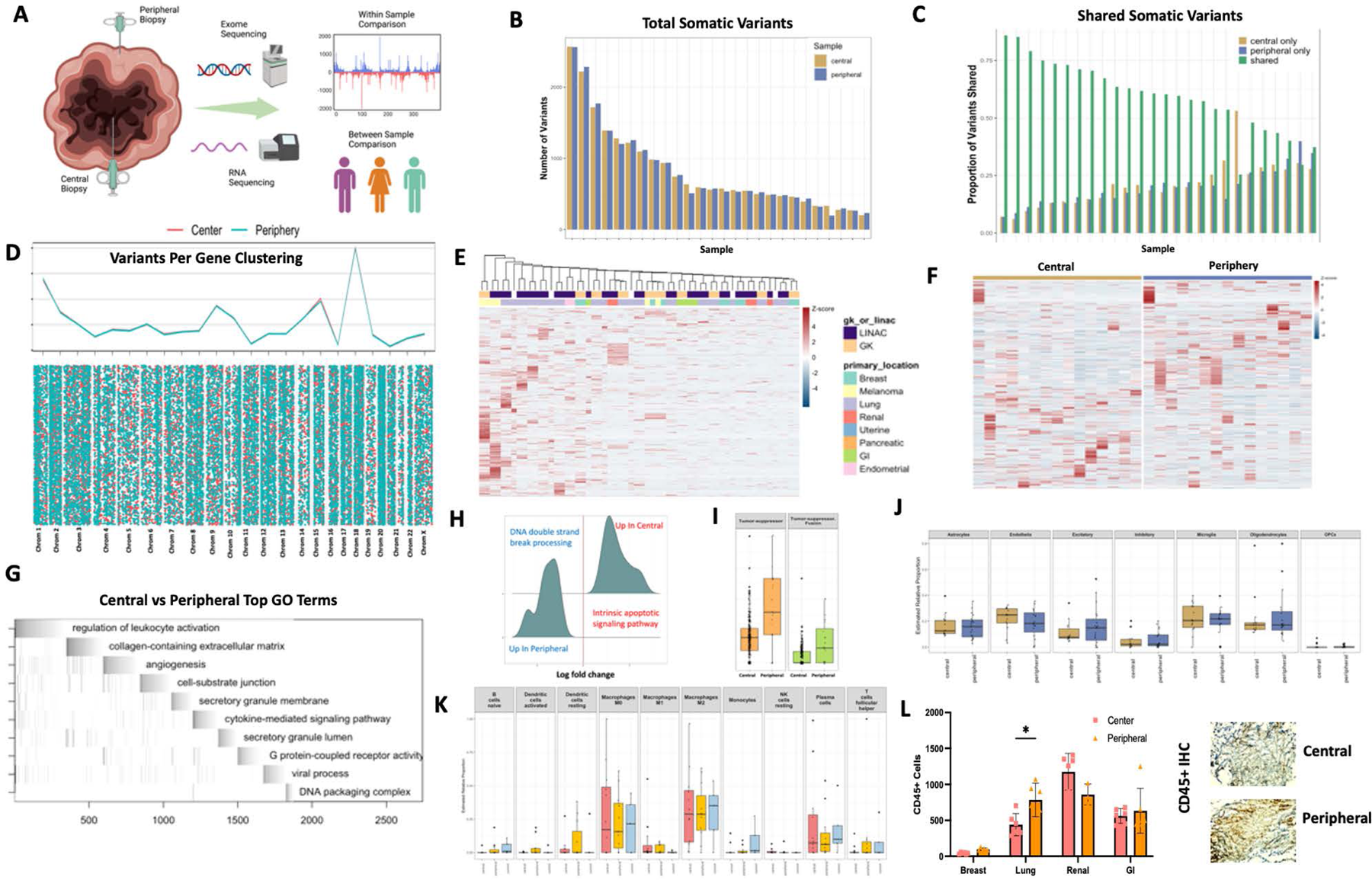
Radiation dosing fall-off along peripheral edges of brain metastasis induces DNA damage repair and enrichment of cellular growth genes. **A)** Schematic depicting isolation of central and peripheral tumor samples during surgery post SRS with comparisons being made across and between tumor types. **B)** Quantification of total somatic variants detected among central and peripheral biopsy locations. **C)** Quantification of shared or unique somatic variants detected among central and peripheral biopsy locations. **D)** Chromosome level visualization of variants from central or peripheral biopsy locations summed (top) or individualized (below). **E)** Heatmap visualization of variants per gene per sample among all central and peripheral biopsy locations across primary tumor location and SRS delivery modality. **F)** Heatmap of differential isoform enrichment analyses conducted by DIFFUSE across central and peripheral biopsy locations. **G)** ALLEZ GSEA waterfall plot with previous term exclusion conducted on differentially expressed isoforms between central and peripheral biopsy locations. **H)** GSEA of differentially expressed genes using ClusterProfiler between central and peripheral biopsy locations restricted to terms included in DNA damage or repair. **I)** Quantification of mutations on genes annotated to be tumor suppressors across central and peripheral biopsy locations. **J)** FARDEEP deconvolution analysis using brain cell type references conducted on central and peripheral biopsy locations. **K)** FARDEEP deconvolution analysis using immune cell type references conducted on central and peripheral biopsy locations. **L)** In-vitro visualization of CD45+ cell invasion using IHC on central and peripheral tumor biopsy locations. Statistical comparisons between groups were conducted with ANOVA with Bonferroni correction (L). Differential expression analysis was done within DESeq2, ALLEZ, ClusterProfiler, and DIFFUSE using adjusted p-value < 0.05 (E, F, G, H, I). * p< 0.05

### Radiation delivery modality and primary tumor location influence genetic signatures and relevant DNA damage measured in brain metastases

Currently, an SRS dose can be delivered to patients through either LINAC or GK treatment modalities^6, 7^. Both systems utilize a different mechanical delivery process and are routinely used interchangeably depending on the facilities present at the location of care^7^. There are, however, known technical differences present among the two delivery types such as dose conformality, treatment times, fractionation, and single isocenter multitargeting^6, 55^. Currently, no direct randomized controlled trial comparisons of both modalities utilized in the treatment of BM’s exist, although studies in meningioma point to GK offering lower effective normal brain tissue dosing, while LINAC reduced the overall treatment duration^56^. Furthermore, a multi-institution study in BM comparing the two-modality showed higher incidence of radionecrosis with GK with similar overall survival^6^. In order to provide biologically relevant human data to this comparison we examined the genomic makeup of tumors isolated after both GK and LINAC SRS with 12 (35%) of our patients receiving GK while 22 (65%) received LINAC (Fig 4A). Examination of variants along chromosomal lengths revealed differential variant location across treatment modalities in center samples and a trend for increased variants within peripheral samples treated with LINAC (Fig 4B). In peripheral samples across all tumor types, LINAC and GK DNA damage levels were similar (mean of variants called: GK: 707, LINAC: 776, p < 0.15), however in matched lung tumors peripheral tumor variant levels increased compared to GK (mean of variants called: GK: 449, LINAC: 1017, p = 0.13) (Fig 4C). Differential expression analysis across all GK and LINAC patients revealed 172 differentially expressed genes at p value < 0.05 with FDR < 0.10 (Fig 4D). GK patients were most enriched for NXPE4 and NOX1 (Log2FC: −26.82, −7.83 p <0.0001) while LINAC patients were most enriched for ACTN3, CT83, and MAGEA10 (Log2FC: 22.36, 22.36, 21.79 p < 0.0001). While both modalities enriched broadly for chromosomal abnormalities and cell division processes, top enrichment genes from LINAC samples included a number of cancer testis antigen (CTA) genes. While their function is still not well understood CTA genes may be involved in tumor proliferation and immune response, with a number of antigens being trialed as vaccine candidates due to their limited expression in normal tissues outside of reproductive organs^57–61^. It’s possible that SRS therapy could induce higher volumes of CTA mutations and thus lead to aberrant expression and possibly increased immune system response to tumors, similar to the abscopal effect, although detailed molecular studies will be required to fully understand this complex biology.

**Figure 4:**
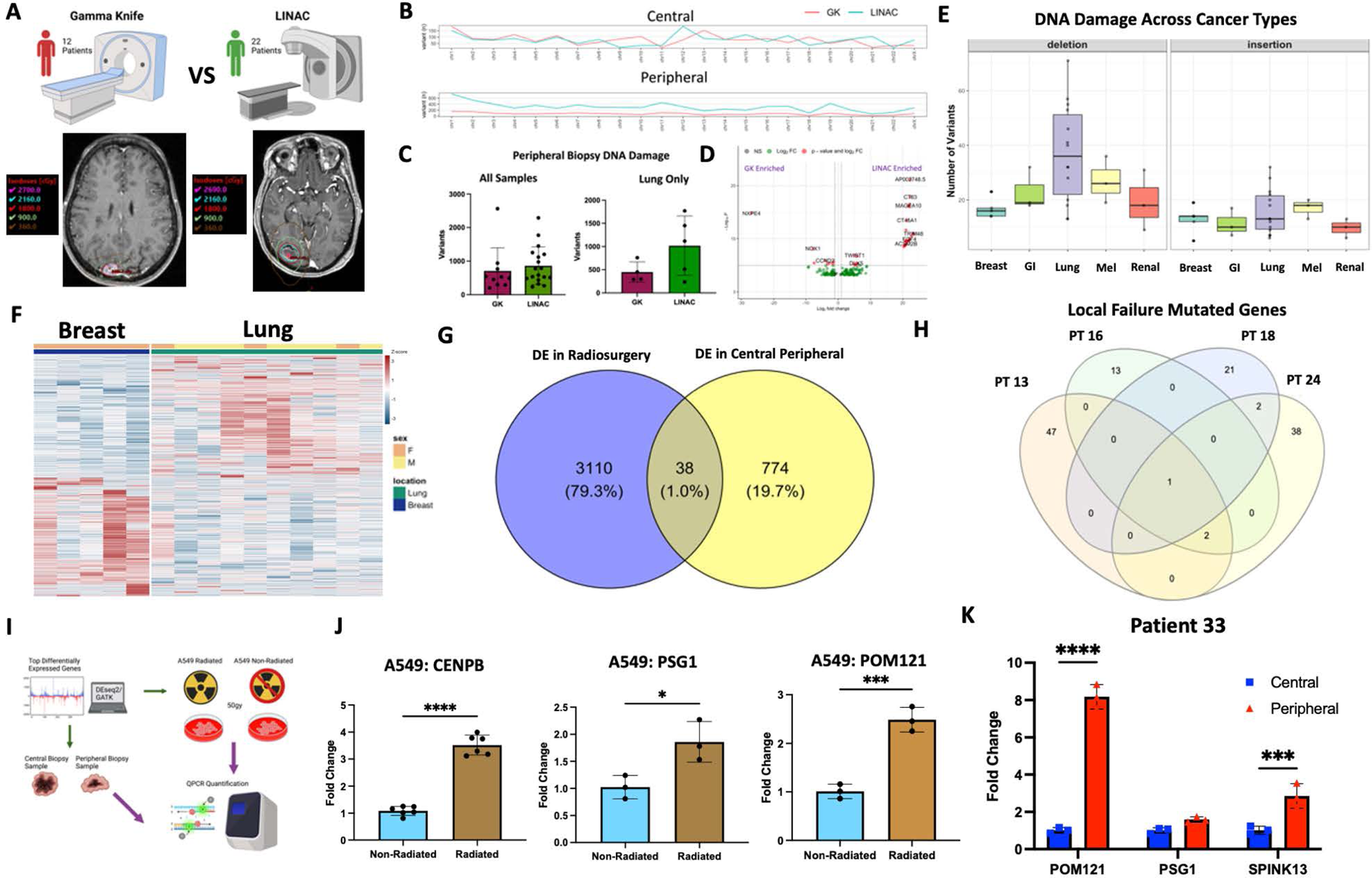
Radiation delivery modality and primary tumor location influence genetic signatures and relevant DNA damage measured in brain metastases. **A)** Schematic depicting comparison between GK and LINAC treated samples with representative images for dose contouring on individual patients. **B)** Chromosome visualization of mutations between GK and LINAC treated samples across central and peripheral biopsy locations. **C)** Quantification of total variants detected among peripheral samples between GK and LINAC SRS delivery. **D)** Volcano plot of DESeq2 determined differentially expressed genes between GK and LINAC treated samples. **E)** Visualization of insertion or deletion variants detected among all primary tumor locations. **F)** Heatmap displaying differentially expressed genes between BM’s of breast of lung primary tumor origin. **G)** Venn diagram **of** differentially expressed genes across radiosurgery v no-radiosurgery comparisons and central and peripheral comparisons. **H)** Venn diagram of high impact mutations across the 4 patients within the clinical trial with documented local failure. **I)** Schematic depicting the in vitro validation of bioinformatic gene hits using QPCR on both isolated peripheral and central biopsy samples as well as radiated A549 primary tumor cells. **J)** QPCR quantification of relative expression of CENPB, PSG1, and POM121, in radiated and non-radiated A549 tumor cells. **K)** QPCR quantification of relative expression of POM121, PSG1, and SPINK13, in isolated central or peripheral tumor biopsies. Statistical comparison between two groups conducted with students T-test (C, J), between more than two groups using ANOVA with Bonferroni correction (K). Differential expression analysis was done within DESeq2, ALLEZ, ClusterProfiler, and DIFFUSE using adjusted p-value < 0.05 (D.F). * p<0.05, *** p<0.001, **** p<0.0001

As current research, as well as our data up to this point has demonstrated, metastases are not all created equal. To this end primary tumor location was examined by visualizing the average frequency of deletions or insertions present across primary tumor types. Results demonstrated that lung BMs carried the most deletions while melanoma samples carried the most insertions (Fig 4E). Breast primary BM’s and lung primary BM’s (the two most common sample types in our study) also exhibited differential expression at the RNA level (1477 genes DE at α < 0.01) (Fig 4F). Examination of genes that were frequently mutated as well as differentially expressed revealed a cohort of 38 genes (1% of our sample results) that fit both categories, interestingly one commonality CENPB, a protein that facilitates centromere formation, occurred in these results as well as was mutated in all of our patient samples that experienced local treatment failure (Fig 4G & E). Clustering of all patient samples based on gene expression utilizing UMAP of defined principal components was also conducted which illustrated within patient similarity but between patient differences with even similar typed tumors failing to reliably cluster together (Sup fig 4A, B & C). To validate the in-silico analysis findings *in vitro* QPCR from either irritated (50gy) A549 primary lung cancer cells or isolated peripheral and central tumor samples was conducted on top hits (Fig 4I). Enrichment for CENPB, PSG1, and POM12 (p < 0.0001, 0.5, 0.01) was confirmed in radiated A549 cells as well as enrichment of POM121 and SPINK13 (p < 0.001, 0.001) within peripheral samples compared to central samples (Fig 4J & K). Mutated status of many upregulated genes (obscurin, mucin, usherin, and homerin) was also visualized indicating enrichment among central samples (Sup Fig 4D, E, F & G). Together these genes all belong to families highly enriched for cell division and replication further confirming that within human samples SRS delivers detectible genomic damage that can disrupt cellular division and proliferation. Furthermore, results from this data indicate a potential biological difference in the damage delivered between different SRS modalities, a finding that certainly requires furthermore detailed clinical study.

## Discussion

As SRS is increasingly being used in the clinical treatment of BM’s an understanding of the genetic underpinnings of the damage it causes is paramount, however, human genetic data from radiated tumors has yet to be examined. In this study we provide the first in human dataset examining the genomic effects of radiation therapy on BM. Consistent with prior studies using *in vitro* human cells^22, 23, 44, 62^ and mouse tumor models^63–65^ significant genomic changes are induced by SRS. Our results clarify that these genetic changes, driven mainly by SNP’s, can be detected in both RNA and DNA across a time series of up to 4 days between dose and resection, although damage likely persists longer. Examination of our results in the context of other large genomic analyses of BMs^14, 15^ that were not treated with radiotherapy identified both common and unique mutations (TP53 commonality, CENPB radiation specific) indicating that biologically radiation therapy provides a unique evolutionary selection pressure, possibly being exploited through the further mutation of tumor suppressor genes.

Although SRS can eliminate lesions, locale failure is documented in 10-20% percent of cases at 1 year post treatment^2, 29, 66^. Because of the challenges of contouring and dosing both larger and irregularly shaped resection cavities as well as concern for surrounding cortical tissue, an aggressive dosing falloff to the periphery of the tumors is often observed. With this study we demonstrate for the first time in humans that clinically delivered SRS doses to both the center and periphery of BM’s have differential genetic effects. This differential effect of radiation induced damage may contribute to both the development of leptomeningeal disease from dissemination of viable tumor cells during surgery and local failure post SRS dosing and clinical resection. Our results provide important biological context currently lacking in the radiobiology field and may be useful when designing future clinical treatments for BM.

Depending on the treatment location and the technology available at the respective treatment center, SRS can be delivered by either LINAC or GK delivery systems. LINAC systems can employ a single isocenter multitarget treatment technique (SMIT) which allows for a more rapid treatment delivery but incurs a small dose conformity penalty^6^. Clinical trials directly comparing between the different delivery systems are currently ongoing, but retrospective analysis of BM patients treated with each modality revealed similar survivals between the cohorts, however, a significant increase in radionecrosis was seen in the GK treatment group^6^. In our cohort we were able to compare genomic sequences from tumors treated with either GK or LINAC systems and confirmed differential genomic signatures and mutation locations across treated patients. This can equip clinicians with direct biological evidence that consideration of the dosing modality should be factored into treatment strategy.

Finally, because our patient population included a variety of primary tumor locations, the study of the effects of SRS across tumor types was possible. Although all tumors that were analyzed are classified as BM’s we found expected similarities and unexpected differences. Consistent with previous literature, radiation induced genomic signatures common with DNA damage such as chromosomal abnormalities, centromere disfunction, nuclear pore formation, which were seen across tumor types. Given radiation is a target agnostic treatment modality similarity between the primary tumor types should be seen, however, specific mutational differences as well as a differential mutational burden following SRS therapy indicates that primary tumor origin may impact radiation response. This has been hypothesized but not yet confirmed in human studies and opens the possibility of primary tumor specific therapeutic augmentations to improve radiation response, such as a radiation sensitizing agent for renal tumor metastasis which displayed a lower mutation burden post SRS. Conversely, metastasis from primary tumors that responded well to SRS treatment, displaying more DNA damage, could be more aggressively treated either within the brain or at the primary site.

Overall, this studies strength comes from the unique human data that is generated, which to date has not yet been collected or studied. Furthermore, the profiling of both DNA and RNA through advances sequencing technology, as well as spatial examination of SRS delivery, allows for a more holistic analysis of the effects of radiation on human cells *in vivo.* The study is however limited by the small number of samples in some primary types, which directly impacts the statistical power to draw conclusions. Lack of germline genomic control samples, and samples from the primary tumors, in our exome analysis also hampers our ability to draw conclusions relating to clonal tracking and evolution causing us to focus on only treatment effect.

In conclusion, with the first comprehensive genomic analysis of radiation effects in human BM we aim to arm clinical providers with biological and genomic data to inform evidence based clinical evaluation of treatments of a tumor type that has become increasingly pervasive in oncological care. Future clinical and biological study of these effects will be necessary in order to understand the effects of treatment both on local and systemic BM control, with the aims of both increasing survival as well as limiting treatment related toxicity and cognitive impacts.

## Methods

### Clinical Trial Enrollment and Patient Selection

NCT03398694 clinical trial protocol was previously published^9^ and adhered to for the duration of this study. Briefly, patients with up to 4 symptomatic brain metastatic lesions were consented and enrolled for pre-operative (1-4 days) SRS. Following delivery of SRS, the lesions were resected with the center and periphery of the lesions being marked and removed during surgical resection. Patients were monitored for 6-month local control as well as leptomeningeal disease status. Follow up visits for patients were scheduled for 1 month post resection as well as every 3 months thereafter for two years (with collection of an imaging sequence). The clinical trial was approved and monitored by the IU Simon Cancer Center institutional review board and data safety monitoring committee.

### Sample Collection and Isolation

During surgical resection of the SRS treated BM’s the center and peripheral portions of the lesion (as determined by the operating surgeon) were collected, removed from the OR, and then flash frozen using liquid nitrogen. Samples were then stored at −80 until isolation of DNA and RNA was able to be completed.

The AllPrep DNA and RNA isolation kit from Quiagen was utilized according to manufactures instructions to obtain viable DNA and RNA from patient samples. Samples were pre-minced with a tissue homogenizer and then run through a Quiagen shredder column to ensure homogenization before proceeding to AllPrep Once obtained, genomic material was QC’d and quantified using a thermo fisher Nanodrop system as well as separately QC’d by sequencing facility using Qubit and Bioanalyzer. Once DNA and RNA were obtained for patients the resulting material was stored at −80 until submission for sequencing. All patients (34) were able to provide some viable genetic material (peripheral/center DNA or RNA), however, due to the nature of the samples being dose with radiation and often necrotic isolation of high-quality RNA proved difficult for some samples, especially within the dosing center of the lesion. Control non-radiated BMs were separately obtained from the Northwestern University clinical tissue bank and were processed in the same manner as above samples.

### Sequencing

All sequencing for samples was performed by Novogene in accordance with company standards and protocols. Briefly, WES was performed using the Aligent Sure Select V6 kit and sequenced with 150bp paired end reads yielding a target of 12GB per sample run. RNA sequencing was performed using poly-A tail enrichment with NEBNext Ultra II RNA kit sequenced with 150bp paired end reads with a total target of 30M reads per sample. At the completion of sequencing raw FASTQ files were delivered to us for downstream bioinformatic analysis. Upon receipt of FASTQ files MutliQC^67^ was utilized to ensure all files passed quality control metrics (detailed in Supp Fig 1).

### WES Analysis

WES analysis for all samples was completed using the well-established GATK pipeline in order to entourage reproducibility of the project^68, 69^. Due to the nature of the clinical trial genomic control from whole blood was not able to be obtained from patients so a panel of normal variant calling approach was utilized. In this approach, documented well in the GATK pipeline, the panel of normal created by broad for hg38 (our reference genome) was utilized for calling of short variants (SNP and Indel) across both our non-radiated controls as well as our SRS treated samples. The rest of the GATK pipeline was followed as standard, with the output VCF files being annotated by SNPEFF^30^. The COSMIC database was also used for functional classification of mutations across and between samples. Quantification of variants and eventual visualization was completed in R v 4.30 using GGPlot, PlotVCF, and pheatmap, hosted in Bioconductor.

### RNA-Seq Analysis

RNA sequencing analysis started with all FASTQ files passing QC via MultiQC (with adaptor trimming being completed when necessary) and began with alignment using STAR^70^. After alignment normalization was completed with RSEM^71^ and the resulting normalized count matrix was supplied to DESeq2 and differential expression was analyzed according to recommended methods by package developer. ALLEZ and ClusterProfiler^72^ were used for GSEA analysis on differentially expressed genes within our samples and FARDEEP^54^ was employed for deconvolutional analyses. For isoform enrichment Diffuse^49^ was utilized after confirming convergence at the .001 level and ensuring model fit parameters using QQ and density plots. Isoform enrichment was considered significant at the α <0.05 level.

### QPCR

To validate in-silico differential expression hits we utilized QCPR to measure gene upregulation (relative to gapdh control) in both our biopsy samples as well as the lung cancer cell line A549. Samples were run on an Adaptive Biotechnologies QPCR machine utilizing SYBR green dye from Bio-Rad. Expression changes were calculated using delta-delta CT and visualized in Graphpad Prism. At least technical triplicates were run for each experiment.

### Immunohistochemistry

Immunohistochemistry staining for CD45+ cells in our peripheral and central samples was completed by the TRIPP pathology lab at UW-Madison Carbone Cancer Center. CD45 antibody staining was validated on non-precious control samples and was then stained on cut sections from our patient biopsy samples. Quantification of CD45+ cell invasion was obtained using IGV as well as verified by two separate and experimentally blinded human counters. Prism was used for visualization of results.

### Statistical Analysis

Throughout the manuscript p <0.05 was considered as statistically significant and is reported using * within the figures. All statistical tests and their results are reported within the results secretion of the manuscript. For multiple comparisons adjusted p-values are reported and utilized for the classification of statistical significance. Differential expression statistics were calculated in DeSeq2 and ALLEZ for their respective analyses and adjusted p-values were used to correct for multiple comparisons. For the comparisons between radiated and non-radiated samples central biopsy location was used to represent radiated samples. Within ALLEZ previous term exclusion was utilized for GSEA comparisons and waterfall plot readouts. For comparison between two groups without multiple comparisons students T-test was utilized while comparison between 2-4 groups was handled with ANOVA using Bonferroni correction for multiple comparisons. Cartoon illustrations were created using Biorender.

### Data Availability

All data and code utilized in this project will be made available publicly upon the publication of the project. Raw and processed sequencing data, as well as analysis code, will be hosted on GEO with supplemental manuscript tables providing all gene lists generated within the downstream analyses.

## Data Availability

All data used for the manuscript will be made available upon publication

---

**Supplementary Figure 1:**
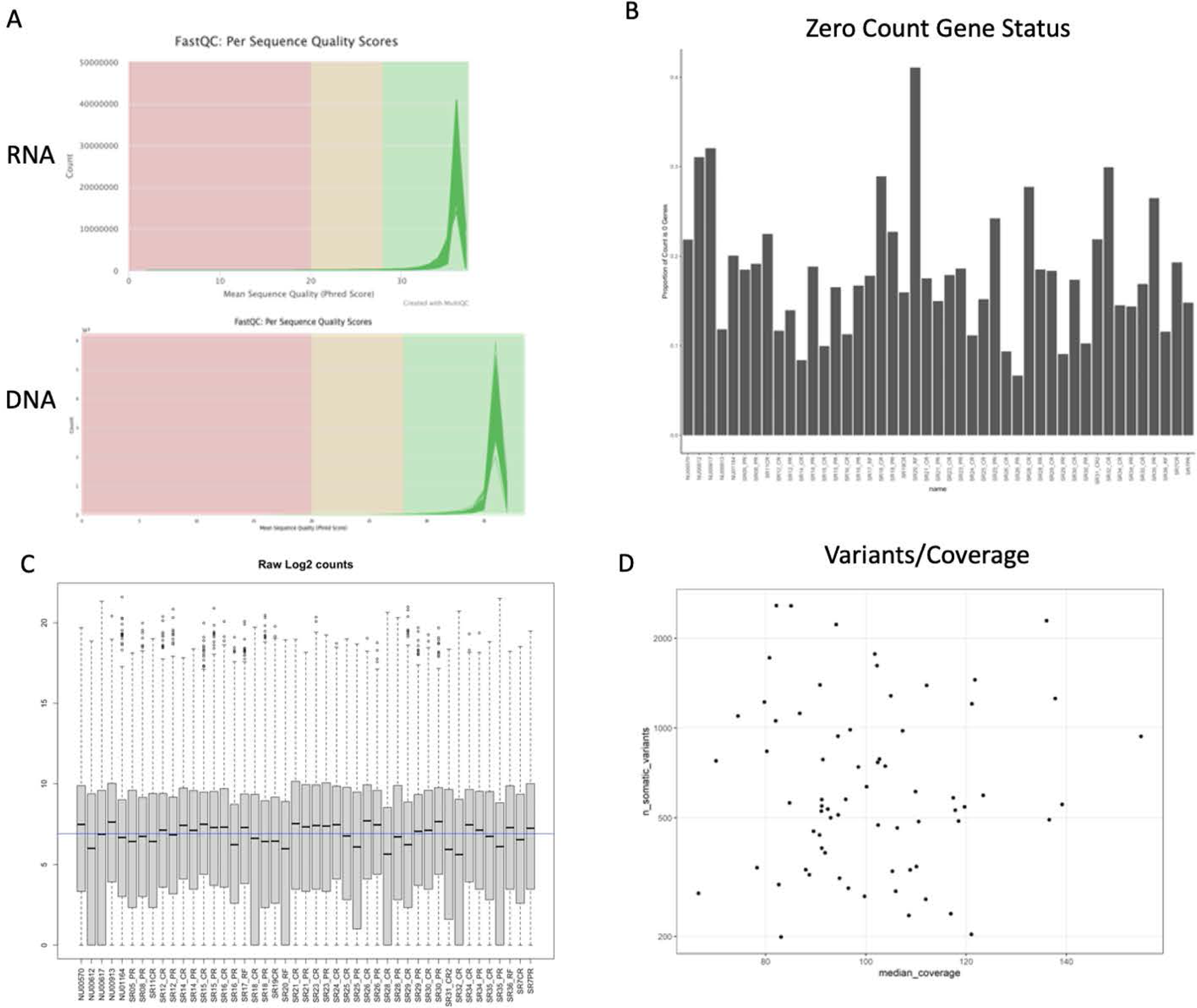
Relevant quality control of sequenced samples. **A)** FASTQC per sequence quality scores across all samples for WES and RNA-Seq. **B)** Proportion of zero counts among RNA-seq of all samples. **C)** RAW log2 counts per sample. **D)** Plot of variants detected per coverage obtained across samples.

**Supplementary Figure 2:**
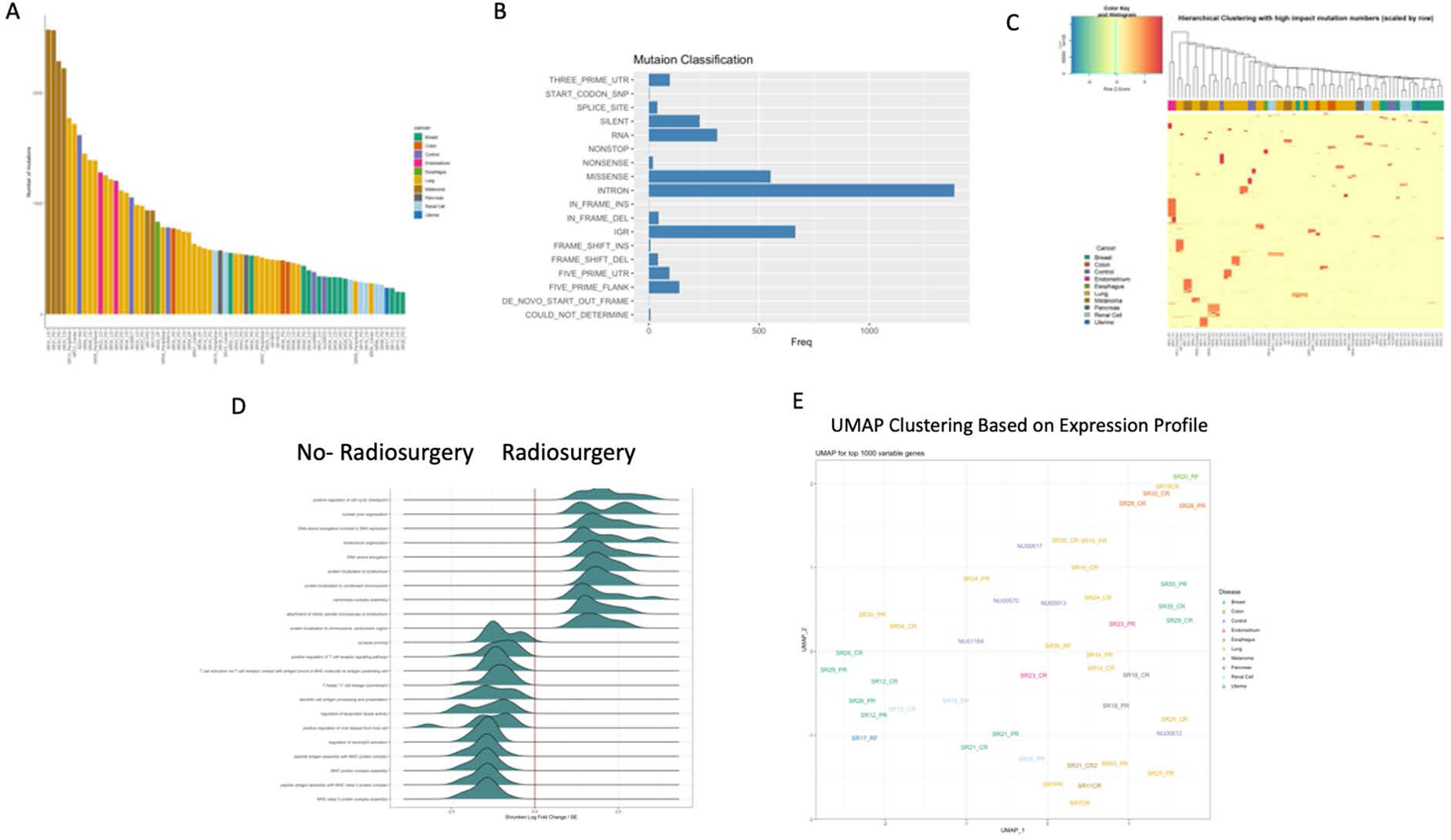
Radiosurgery vs No-Radiosurgery comparisons among all sample types. **A)** Plot of number of variants detected per sample annotated by primary tumor location. **B)** Classification of type of mutation found within samples and its frequency. **C)** Hierarchical clustering applied to variants detected across samples. **D)** ALLEZ GSEA of all significant terms between radiosurgery and no-radiosurgery groups (not restricted to lung primary tumor location). **E)** UMAP clustering applied to top 1000 variable genes and annotated with tumor type.

**Supplementary Figure 3:**
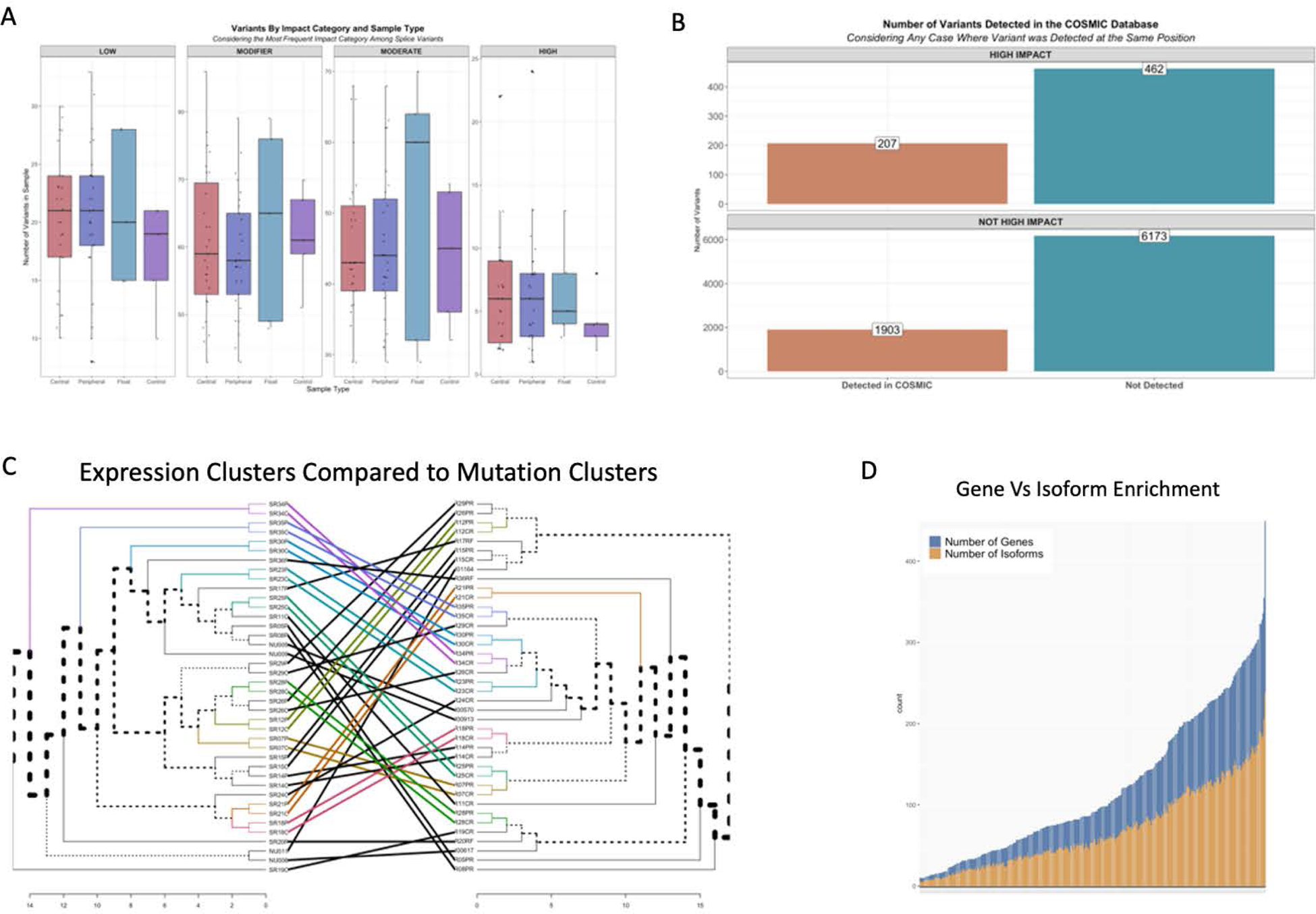
Gene annotation and isoform enrichment between central and peripheral biopsy samples. **A)** Impact of variants called among all sample types with float representing samples where central or peripheral location could not be determined. **B)** Results of annotation of variants using the COSMIC dataset. **C)** Chaos plot of clustering between WES data and RNA-seq data. **D)** Visualization of gene vs isoform enrichment among all samples.

**Supplementary Figure 4:**
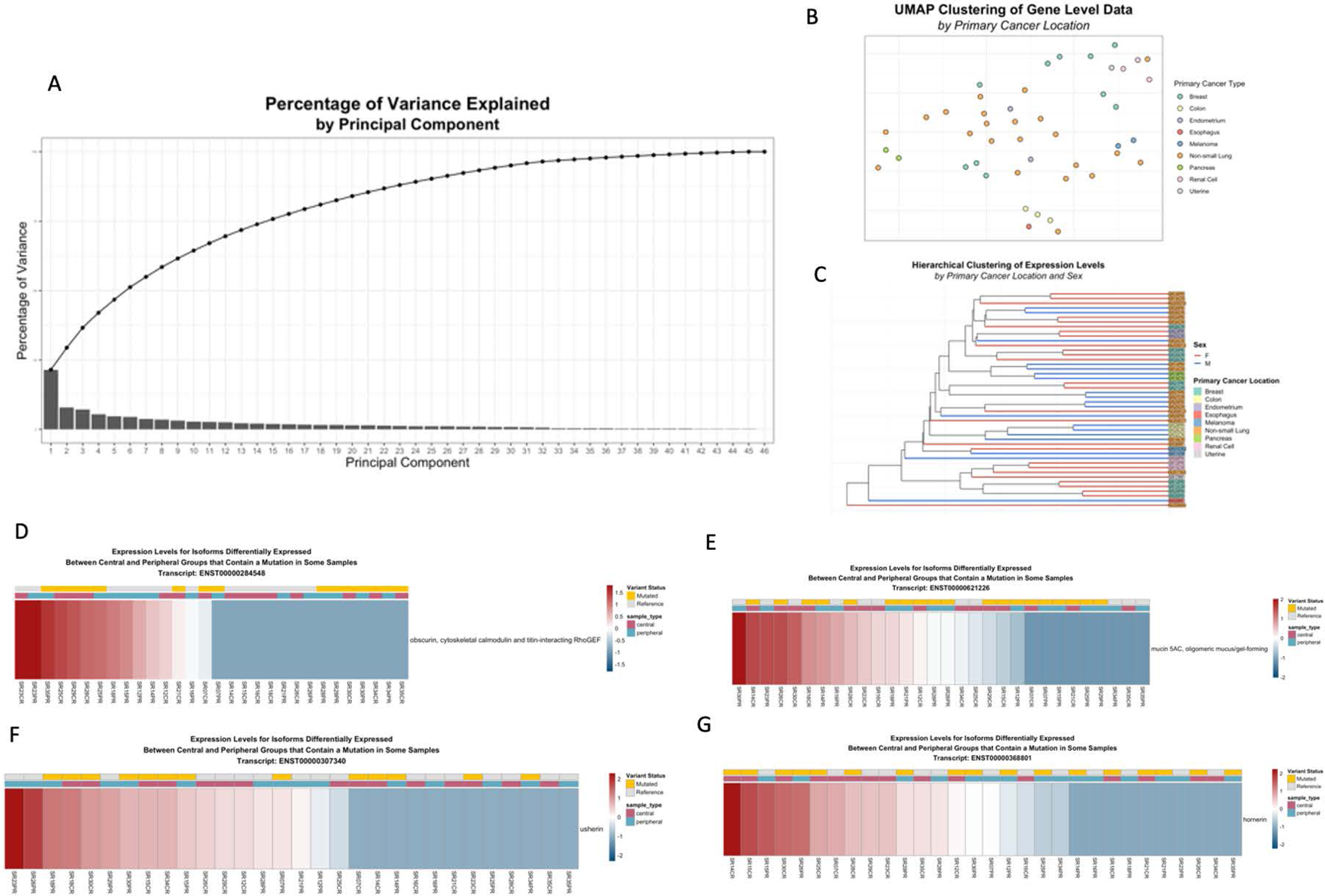
Clustering among all patient samples along with RNA expression levels of genes with detected mutations. **A)** Visualization of principal components used for dimensional reduction clustering. **B/C)** UMAP and hierarchical clustering of samples by tumor primary location and primary location and patient sex, respectively. **D/E/F/G)** Visualization of gene expression levels (RNA-Seq) among genes that were annotated to have a variant (WES).

## Acknowledgements

The authors thank the patients who bravely contributed samples for this research and whom all this work is dedicated to.

## Author Contributions

JMS/QW/ZN/GC conducted bioinformatic analysis, JMS/LZ/XW/AP/LM/VK/NG conducted in-vitro experiments and isolated/prepared samples for sequencing, AA/GP contributed to manuscript formulation and provided sequencing samples and cell lines, NA/JM/CK/ACG/TP/CL/JS/BL/KS/AK/MS/GW provided clinical consultation and/or operated on patients on trial, JMS/MD/CK wrote the manuscript, MD supervised the project.

